# Estimating the health impacts of climate change for policy decision-support: a systematic review of spatial microsimulation methods

**DOI:** 10.1101/2025.10.03.25337089

**Authors:** Ariel A. Brunn, Roberto Picetti, Lauren Ferguson, Francis Ruiz, Petra Meier, Rosemary Green, James Milner

## Abstract

Spatial microsimulation models have recently emerged as a new method to quantify health impacts associated with climate change for policy decision-support. These individual-based methods, previously used in tax and health policy planning, have been adapted by combining climate data with exposure-response associations to estimate the distributional health impacts attributable to climate hazards using synthetic populations. To evaluate their methodological characteristics, we conducted a systematic review of the literature. We searched five electronic databases, Google Scholar and the International Journal of Microsimulation, and screened 762 articles to reach a final study set of seven articles. Most models simulated populations based in high income countries (n=5) and applied dynamic methods to forecast future health outcomes (n=5). Multiple diverse climate-health pathways of impact were investigated, ranging from heatwave mortality to air pollution-induced cardiovascular outcomes, to climate-sensitive infectious disease occurrence. Baseline and projected spatial climate data was mapped to individuals in city, state, or regional-level synthetic populations to allocate personal hazard exposure. Most models included socio-economic and demographic attributes (n=6) to integrate vulnerabilities for burden assessments in marginalised groups such as children, women, and the elderly. Climate policies mainly focused on mitigation and simulated future emissions scenarios (n=5), or policy mixes (n=1); one study tested an incremental adaptation intervention. Methods to enhance decision-support among alternative policy options such as economic evaluation (n=2) or stakeholder engagement (n=3) were under-represented. All models acknowledged uncertainty of parameters, and most reported uncertainty analyses (n=5). High data needs may limit accessibility of these methods in some contexts, however options to build on existing models and improve data and computing power access could overcome these challenges. This systematic review documents this evolving, state of the art application of microsimulation and finds a promising and versatile quantitative tool for health impact assessments and climate policy decision-support.

## Introduction

The concurrent rise of big data, open-access publishing, and machine learning has accelerated methodological development of complex data-driven mathematical models to support decision-makers. Microsimulation (MSM) is one such method established to optimise the policy development and design process through comparative assessment of interventions and applied to a representative “synthetic” population generated using survey and census data of existing populations (Rutter et al. 2011; Schofield et al. 2018; Lomax 2022). This modelling method simulates individuals using static or dynamic processes and spatial methods allocate individuals into geographical zones across cities, states, or countries, using deterministic or probabilistic methods (Lovelace & Dumont 2016). Its métier of capturing distributional population impacts has long supported its application for government policy planning including taxation and health, as envisioned by Guy H. Orcutt in 1957 (Ballas et al. 2013; Orcutt 2007). More recently, these individual-based methods have drawn the attention of climate and health scientists investigating the social inequities of climate change.

These inequities arise from uneven exposure to hazards such as heatwaves, floods, droughts, storms and air pollutants, the latter of which share sources with carbon emissions and whose concentration, distribution, and impacts on radiative forcing are themselves influenced by temperature, rainfall and wind patterns (Orru et al. 2017). Exposure to these climate hazards and individual coping ability is mediated by socio-economic and demographic factors, leading to differential risks within populations. To better understand and quantify the range of health consequences associated with climate hazards, methods of statistical attribution have been developed drawing largely on environmental impact assessment, with more recent advances aimed at detecting signals from long-term (decadal) changes in climate trends (Ebi et al. 2020; Carlson et al. 2022). This increasingly quantitative and health-focused approach strengthens climate policy efforts by foregrounding the human costs of climate change - including stark differences in risk - into discussions of common responsibility and fairness of climate mitigation and is particularly important to low-and-middle-income countries (LMICs), where poor governance, limited financing, and insufficient health infrastructure exacerbate population disparities.

Most current climate-health modelling methods only weakly address differential vulnerabilities associated with climate exposures. Integrated assessment models (IAMs), which have substantially advanced modelling efforts by combining science and policy elements and more recently incorporated health impacts (Weber et al. 2023), nonetheless mainly rely on aggregated data to reduce uncertainties associated with forecasting future climate scenarios (van Beek et al. 2020; Jafino et al. 2021). Since aggregative methods obscure individual heterogeneity occurring at different timescales and geographies, the full spectrum of exposure risks and vulnerabilities cannot be considered. This masking of differences in how people experience climate change reinforces existing injustices and engenders others, and when used as the basis for policy development, increases the likelihood of uneven benefits for some groups and unintentional harm to others (Thomas et al. 2019; Newell et al. 2021).

In the parlance of climate policy, distributive justice provides a framework for the fair and just allocation of costs and benefits among individuals exposed to climate change and the value system under which this allocation is made (Vermunt and Tornblom 1996, Jafino et al. 2021). Several features of MSM lend themselves to considerations of justice. For instance, spatial MSM methods permits geographic variation of climate hazards to be matched to geo-located individuals at a high resolution to better capture the range of personal exposures that different people experience (Lovelace & Dumont 2016). Further, the process of simulating individuals through the creation of a ‘close-to-reality’ synthetic population offers improved granularity of socio-economic attributes that drive climate-health vulnerabilities in sub-populations, uncovering the non-random spread of inequities (Kopasker et al. 2023). Additionally, policy testing can be applied to sub-populations disaggregated by a number of variables, enabling the exploration and comparison of impacts and costs across diverse social circumstances and accordingly, form a basis to refine and optimise climate policy design for greatest benefit. Collectively, these features form the basis of methods which may be able to generate more specific estimates of population health burdens based on individual exposure risks without need for simplifying assumptions of homogeneity, and when combined with sector-specific levers, can inform a targeted policy response.

As progress slows on greenhouse gas (GHG) emission targets under the Paris Agreement, attention is turning to the policymaking process itself and efforts to improve its effectiveness and scope (OECD 2024). Acknowledgement of the limitations of aggregate simulation methods and the emergence of individual-based models for climate policy has driven interest in MSM as a tool to systematically and quantitatively assess the impacts on population health of climate change for policy decision-support. Recent reviews of MSM applications for mental health outcomes and food policy simulations highlight a growing interest in tailoring and enhancing its methods for subject-specific use through methodological appraisal (Merten et al. 2022; de Oliveira et al. 2024). This systematic review therefore aims to provide an overview of MSM methods and data sources to assess health outcomes of climate hazard exposures by describing characteristics of existing examples in the peer-reviewed literature and to assess the value and credibility of these models as decision-support tools.

Review objectives

1. To describe characteristics of MSM applied to climate-health impact assessment, including methods used for synthetic population generation and exposure-response associations.
2. To assess the scope of climate policies applied in the included study set of peer-reviewed literature.
3. To appraise the validity, scope and current challenges of MSM methods and to guide future model development for climate-health impact assessment and policy testing.

## Methods

An a priori protocol was developed using the Preferred Reporting Items for Systematic Reviews and Meta-Analyses for Protocols (PRISMA-P; Shamseer et al. 2015) to reduce sources of bias in the review and registered on the Open Science Framework database (Brunn 2025). To identify relevant articles, some definitions were necessary. Microsimulation was defined as an individual-based method that simulates interactions between an individual and its environment. A spatial approach involves “the analysis of individual-level phenomena over geographical space [and] the creation, analysis and modelling of spatial microdata” (Lovelace & Dumont 2016) and typically applies deterministic reweighting methods such as iterative proportional fitting (IPF) or probabilistic methods such as simulated annealing (Tanton 2014; Lomax 2022; Wu et al. 2022).

### Search Strategy

The search strategy revolved around three subjects, namely, ‘microsimulation’, ‘health outcomes’, and ‘climate change’ and synonyms used as search terms were combined with Boolean operators to conduct the initial electronic database searches. The search string was first formulated through an iterative process in two electronic databases, MEDLINE and Web of Science, to refine the search terms and to identify a list of known publications to internally validate the final database search. The search string is available in the protocol (S3, supplementary materials).

Five electronic databases, MEDLINE, Embase, Scopus, EconLit and Web of Science, were searched for peer-reviewed published literature on January 15, 2025. The same search string was applied in the Google Scholar search engine and the International Journal of Microsimulation was hand-searched for additional relevant articles that may have been missed in the database search. This review focused on peer-reviewed literature and therefore grey literature was not included (Table 1). Abstracts in languages other than English were excluded; recent reviews suggest that MSM research outputs overwhelmingly come from the USA, United Kingdom and Australia (Schofield et al. 2018; Mertens et al. 2022; de Oliveira et al. 2024). Nonetheless, no geographic limitations were placed and studies published in any location or featuring a synthetic population from any global region were included. Given the historic adoption of the Paris Agreement by 196 Parties and concomitant requirements for countries to commit to climate action planning via their Nationally Determined Contributions (NDC), only studies published since 2015 were included. In addition, we included air pollution studies in our inclusion criteria as air pollution often shares common sources with fossil fuels. However, it is not always clear whether the primary target of a given policy is air pollution or climate mitigation. We therefore included studies where the objective to reduce air pollution would also have climate benefits and was either measured or discussed by the authors.

**Table 1.** Eligibility criteria.

| Criteria | Inclusion Criteria |
| --- | --- |
| Publication Year | 2015 - 2025 |
| Location of Study | Any location accepted |
| Language | English only |
| Study type | Spatial microsimulation studies only |
| Publication type | Peer-reviewed literature only; full-text must be available |
| Population | Any synthetic individual-based population |
| Exposure | Any climate variable such as ambient temperature, precipitation, particulate matter etc |
| Comparator | Any population comparators (absence also accepted) |
| Outcomes | Population health outcomes expressed as a disease burden estimates, e.g. mortality, morbidity, disability-adjusted-life-years (DALYs), quality-adjusted-life-years (QALYs) and clearly articulated climate-relevant policies which included climate emissions scenarios, climate targets, or applications of specific interventions |

This review is wholly concerned with MSM methods. Though there is overlap with agent-based models (ABM), we distinguish ABM as models that focus on simulating behavioural interactions among and between individuals and may involve smaller populations due to increased complexity and computational power needs (Lovelace & Dumont 2016). In addition, integration of ABMs and IAMs have already been subjects of recent reviews (such as Lamperti et al. 2019) and therefore these were considered beyond the scope of this work.

### Data Selection and Management

A reference management tool, Rayyan, was used for de-duplication and screening of articles (Ouzzani et al. 2016). Citations underwent a three-stage screening process beginning with a title screen conducted by a single independent reviewer to remove excluded study designs e.g. “Systematic Review”, or ineligible study populations where this was listed in the title (e.g. models of animal or plant species). Abstracts were then screened by two independent reviewers and advanced to full-text screening according to pre-determined eligibility criteria; full-text articles were similarly screened in duplicate with reasons documented for study exclusion.

### Data Extraction

A custom data extraction template was developed in Microsoft Excel. Three broad categories of data were extracted related to article meta-data, model specifications, and policy scenarios. Model specifications included type of climate exposure, health outcomes simulated, method of synthetic population construction, model methods, exposure-response relationships and data source, simulation projection period, methods of uncertainty analysis and of validation, and author-assessed limitations. Data was extracted on the type of policy scenarios simulated and rationale given, net effects of modelled policies on baseline outcomes (i.e. whether policies positively or negatively impact outcomes) and a summary of any economic methods used. This manuscript aims to describe and appraise model methods and therefore data extracted on net effects of policies on health outcomes will not be further included, they will instead be made available in a subsequent mini-review.

### Critical Appraisal

To evaluate the integrity of included models for climate-health policy evaluation, a custom critical appraisal tool was adapted from existing evaluation criteria for climate mitigation modelling of health outcomes (Picetti et al. 2023; Hess et al. 2020) as no published or validated topic-relevant appraisal tool was otherwise available. The tool was divided into three components briefly summarised as: 1) validation of model methods and parameters, including assessment criteria for reporting of key model assumptions, data sources, sources of uncertainty, and validation of generated estimates; 2) credibility of policy simulations which considered author acknowledgement of achievability of simulated scenarios, potential for scale up, and model limitations; and 3) model reproducibility. Each study was assessed by two independent reviewers using the tool to score evaluation criteria on a binary scale; certain criteria could also receive a partial score (0.5). The complete assessment criteria and scoring framework are included in the Supplementary Materials (S2). Aggregate scores for each study were subsequently compiled and visualised for comparison.

### Protocol Deviation

One study prompted a deviation from the protocol. The original protocol required inclusion of spatial methods however, we accepted one study that did not report their use (Stephen & Barnett 2017). Its inclusion was justified on the basis that it was the earliest example of an MSM study that also met all other inclusion criteria and that it highlights the rapid evolution over the last decade through trial and error into a common methodological framework. Spatial methods were not included in the screening criteria, so no bias is anticipated.

## Results

The electronic database search yielded 1,155 records in total for screening (Figure 1), including twenty abstracts from Google Scholar and five from the International Journal of Microsimulation. Abstract screening returned 30 articles for full-text review. At the full-text stage, studies were excluded on the basis that they did not include a simulated climate policy (n=3), lacked a defined health outcome (n=3) or had no climate exposure (n=6), others were evidently the wrong model type (n=7), and one study did not report the construction of a synthetic population. A total of seven studies were therefore included in the final study set of the review.

**Figure 1.**
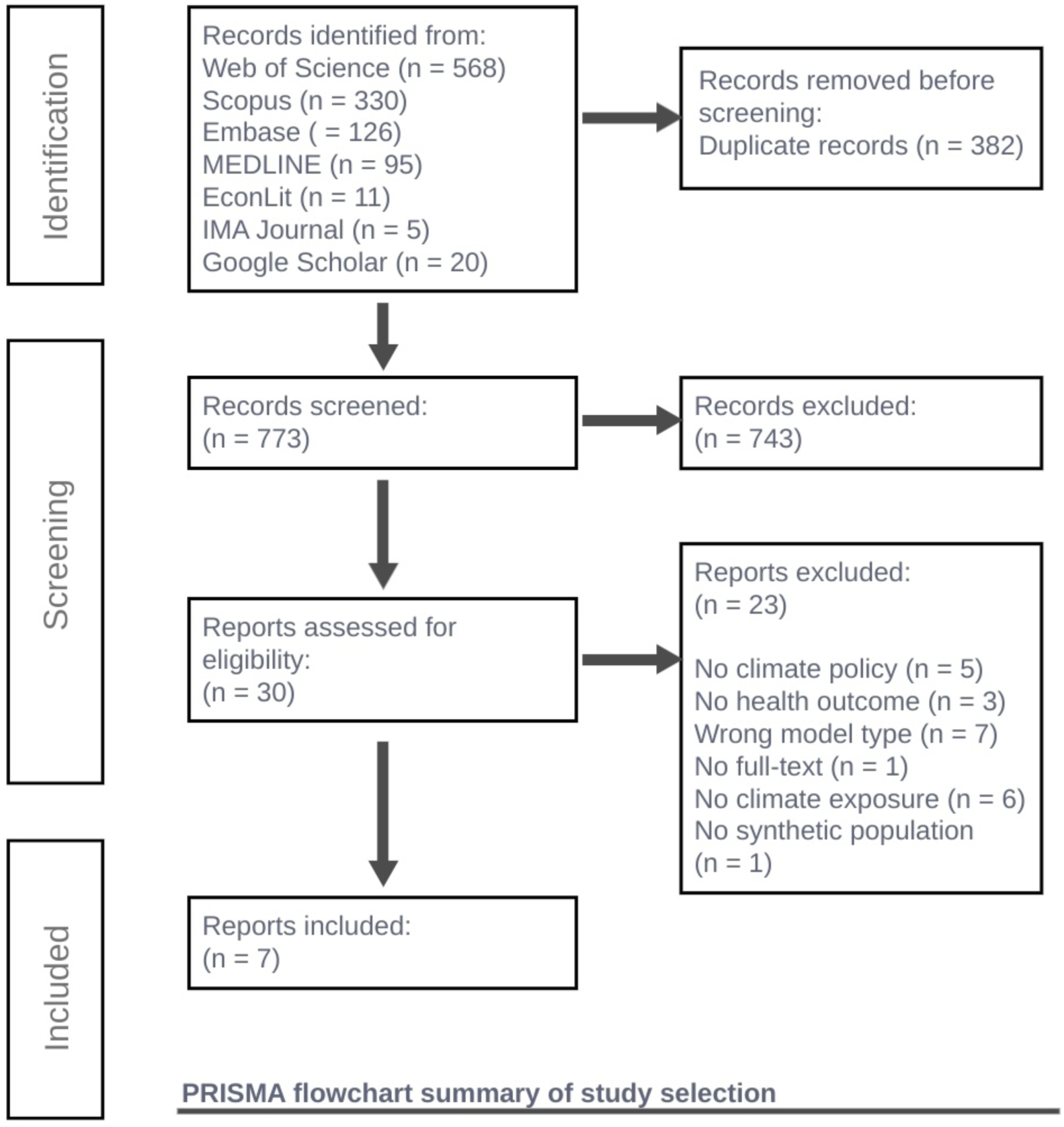
PRISMA Flowchart.

### Article Meta-Data

MSM applied to climate-health impact assessments have appeared in the peer-reviewed literature since 2017 and are published almost equally in either environment or health science journals (Table 2). Most models in the study set simulated populations in high income countries (n=5); there were no studies that met eligibility criteria and that strictly focused on Africa or the Americas. Populations based in LMICs, including China and India, were represented in three studies, however LMIC-based co-author institutions were exclusively from China. Three studies simulated a whole population; the remainder compared population cohorts from a single country (Zeng et al. 2022; Dimitrova et al. 2022), region (Marvuglia et al. 2020), or worldwide sample (Weyant et al. 2018).

**Table 2.**
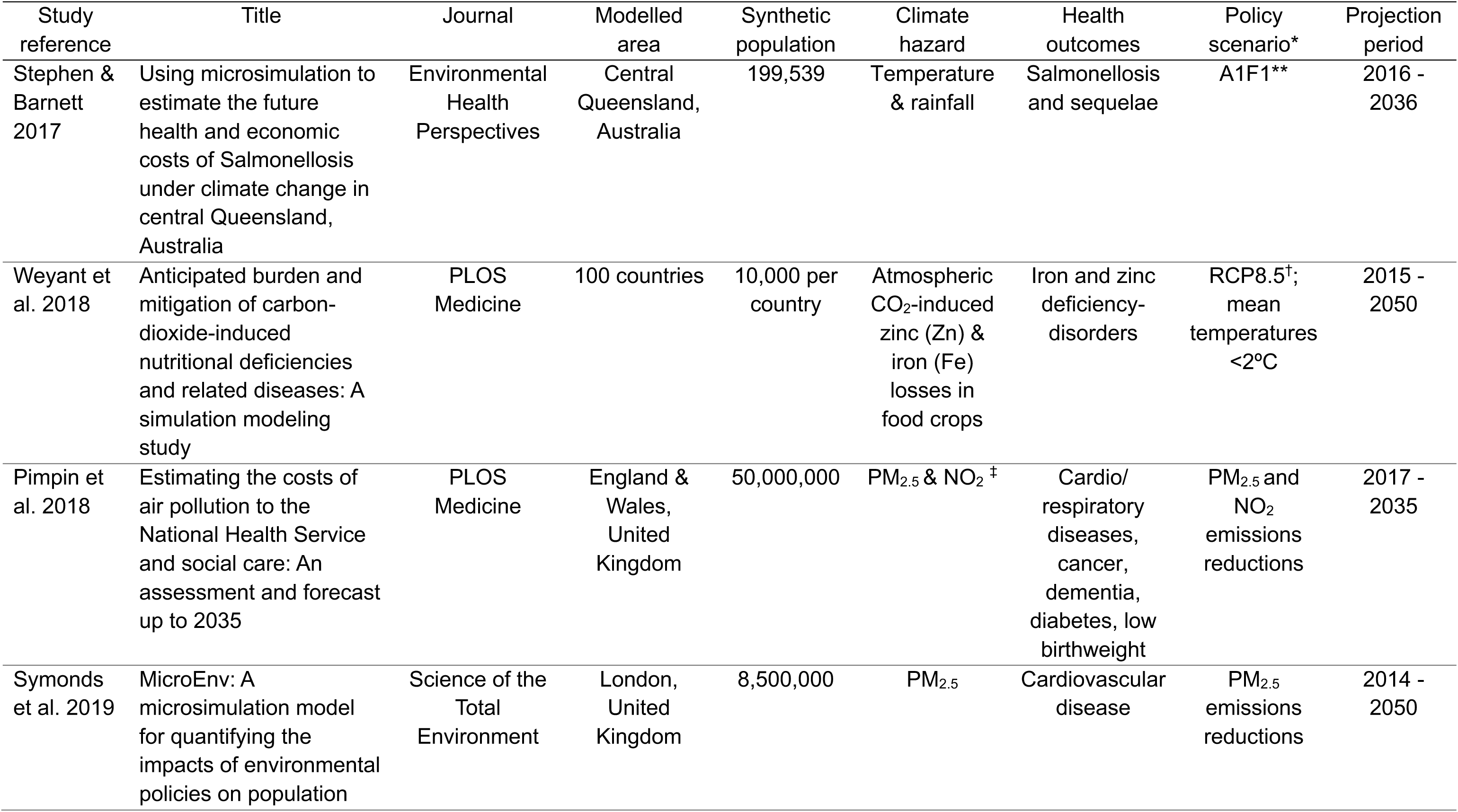
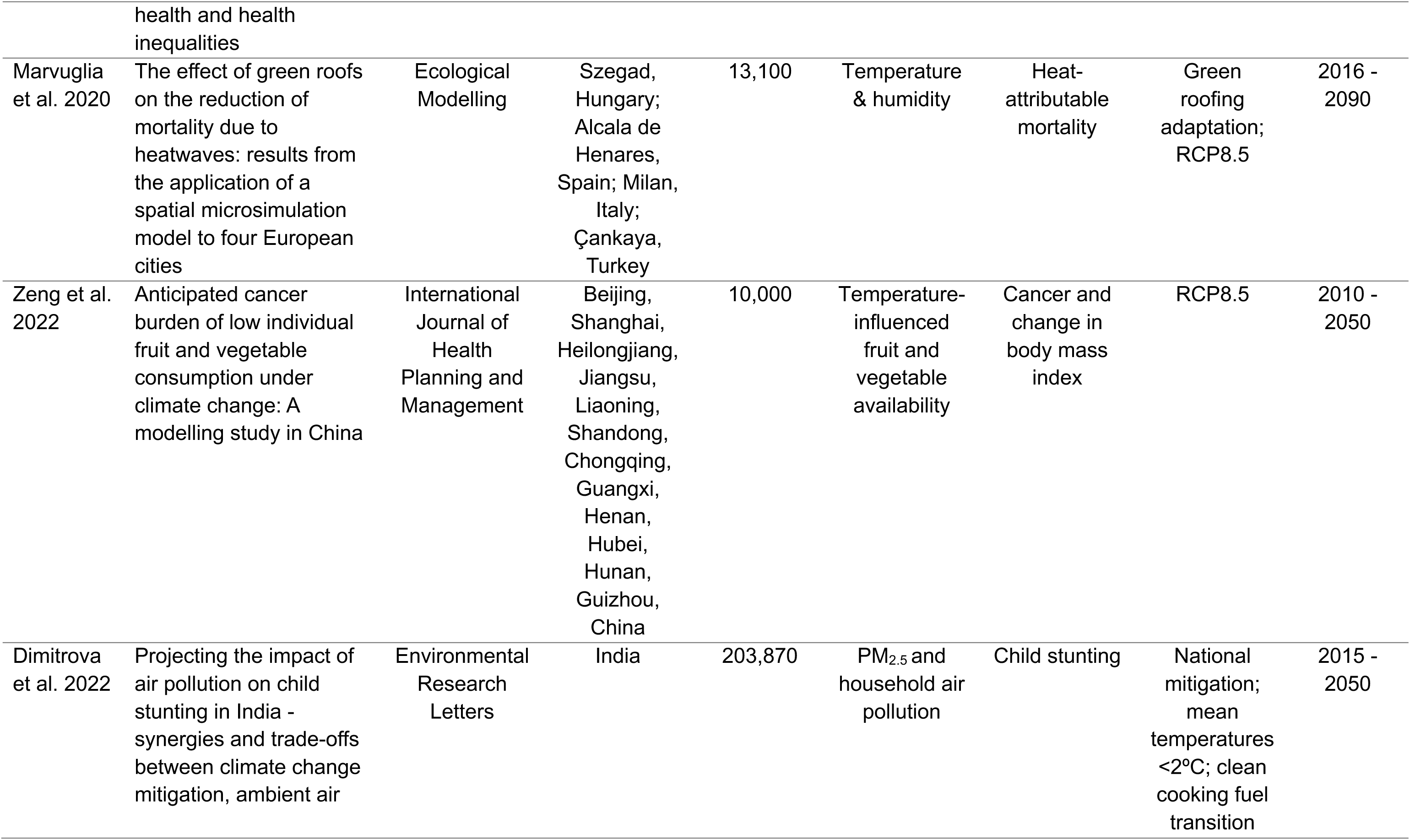
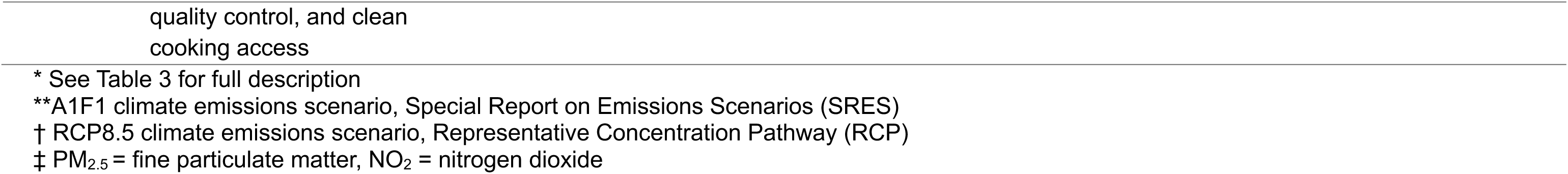
Summary of articles.

### Model methods and population construction

More than half of included studies used dynamic simulation methods to track transitions in individual health states as well as demographic change, apply projections of climate exposures into the future, and incorporate temporal policy effects such as lags (Stephen & Barnett 2017; Weyant et al. 2018; Pimpin et al. 2018; Symonds et al. 2019; Zeng et al. 2022). Two models applied static methods, whereby health impacts were projected at fixed time-points in the future via manual updates of climate input data (Marvuglia et al. 2018; Dimitrova et al. 2022). Projections of health impacts spanned 19-74 years, however most models forecast outcomes up to 2050 (range: 2036, 2090).

Included studies rarely described synthetic population construction methods using conventional spatial MSM methods such as iterative proportional fitting or simulated annealing. Dimitrova et al. reported using reweighting methods in their model of an age-cohort population in India (2022), however many authors only included brief descriptions of population simulation techniques, data sources, or reported the use of population generation through specific software tools such as NetLogo.

The ability to address differential vulnerabilities is a key strength of individual-based methods. Health burden estimations were demographically disaggregated by sex (Stephen & Barnett 2017; Symonds et al. 2019; Weyant et al. 2018; Dimitrova et al. 2022) and age, with some studies highlighting specific groups, such as children under five (Weyant et al. 2018; Dimitrova et al. 2022), the elderly (65+ years, Marvuglia et al. 2020), and working-age people (15 – 64 years, Symonds et al. 2019). Socio-economic attributes varied but included social deprivation (Symonds et al. 2019), maternal education, household income, population caste (Dimitrova et al. 2022), and regional economic development status (Zeng et al. 2022). One study did not report disaggregated health burden estimates (Pimpin et al. 2018).

Most models were coded using R software, with custom packages referenced such as MicSim (Zinn 2024). Three separate studies reported use of either Python, C++, or NetLogo. Two of the largest dynamic population models also acknowledged use of high-performance computing (HPC) services (Stephen & Barnett 2017; Symonds et al. 2018), however, a dynamic model of 50M people in England and Wales did not report the use of parallel processing (Pimpin et al. 2018). Simulations of more populous countries did not necessarily encourage more ambitious population construction – rather, models for China and India used relatively small population samples of 10,000 and 203,870 individuals respectively (Zeng et al. 2022; Dimitrova et al. 2022).

### Exposures and outcomes

Climate exposures comprised air pollutants (PM_2.5_, NO_2_), ambient temperature, rainfall, dewpoint, and carbon dioxide concentrations; the influence of the latter exposure was incorporated through an indirect pathway impacting food crop nutrient availability (Weyant et al. 2018). While most studies centred climate hazard analysis on a single exposure, a model by Stephen & Barnett examined the effect of temperature and precipitation on lagged Salmonellosis incidence (2017) and another studied the apparent temperature or ‘wet-bulb’ temperature estimated based on temperature and humidity (dewpoint) projections (Marvuglia et al. 2020). Baseline climate data sources encompassed historical meteorological data from local weather stations as well as modelled air pollution data sets, while projections were mainly obtained from Coupled Model Intercomparison Projects (CMIP) outputs, or from IAMS, such as projected effects of climate on food crop availability from the agricultural partial equilibrium model, IMPACT 3.3, and emissions for ambient air pollution and clean cooking fuels projected by the Greenhouse Gas Air Pollution Interaction and Synergies (GAINS) and Access household fuel-choice modules of the MESSAGE-GLOBIUM IAM.

Spatial methods were applied to allocate individuals into geographic zones as well as to link climate hazards to geolocated individuals. Matching exposures to individuals or small areas was primarily conducted by linking spatial coordinates using GIS methods (Pimpin et al. 2018; Marvuglia et al. 2020; Symonds et al. 2019; Dimitrova et al. 2022). Two studies applied Monte Carlo sampling of area-specific probability distributions of exposure data (Weyant et al. 2018; Zeng et al. 2022). The spatial resolution of climate data was generally high, ranging from 100m^2^ to 5km^2^ though in two studies historical weather station data was averaged across the full study area (Stephen & Barnett 2017; Marvuglia et al. 2020). Exposure-response relationships were sourced from published literature (Pimpin et al. 2018, Symonds et al. 2019, Marvuglia et al. 2020), the Global Burden of Disease database (Zeng et al. 2022, Weyant et al. 2018) or estimated through regression analysis (Stephen & Barnett 2017, Dimitrova et al. 2022).

While the most common health outcomes investigated were non-communicable diseases such as cardiovascular, respiratory diseases, or dietary deficiencies, two studies measured climate-sensitive infectious diseases (CSID) - salmonellosis and malaria (Stephen & Barnett 2017; Weyant et al. 2018). In the former CSID outcome, a time lag of 21 days was also incorporated to account for *Salmonella* transmission through food chains. Mental health outcomes were not assessed in any model. Standardised health metrics, necessary to provide comparable and generalisable results, were reported in all studies (Table S1, Supplementary Materials).

Both direct and indirect pathways of climate impact were simulated within the study set. The former pathway included heat-attributable mortality and the influence of ambient temperature and precipitation on enteric disease incidence. Indirect pathways were often implemented by coupling sub-models together in which the results of an initial model would be fed into the second (usually the MSM) model; this was common in investigations of the effect of climate change on food/nutrient availability and its associated health outcomes. All dynamic models used probabilistic methods to run health state transitions, including Bernoulli trials (Symonds et al. 2018), Monte Carlo sampling (Weyant et al. 2018; Pimpin et al. 2018; Zeng et al. 2022), and transition rates derived from probability distributions (Stephen & Barnett 2017; Marvuglia et al. 2020).

### Climate policies

Policies varied in their form and objective within the study set (Table 3). Five models applied global climate emissions scenarios using Representative Concentration Pathways (RCPs) or Special Report on Emissions Scenarios (SRES) projections to compare the burden of health outcomes under future climate trajectories. Other models forecast health reductions achieved resulting from specific climate targets, such as keeping mean global temperatures below 2°C. One study compared nutritional health impacts under low and high projected atmospheric CO_2_ concentrations corresponding to low- and high-radiative forcing scenarios (Weyant et al. 2018). Two studies simulated the health effects of air pollution emissions reduction targets for PM_2.5_ and NO_2_ referencing parallels between climate mitigation actions aimed at fossil fuel-based sectors, such as transport and industry, and air pollution-associated health co-benefits (Pimpin et al. 2018; Symonds et al. 2019).

**Table 3.** Description of policy simulations and stakeholder engagement.

| Study reference | Policy scenario | Action | Details | Stakeholder engagement |
| --- | --- | --- | --- | --- |
| Intervention-based policies |  |  |  |  |
| Marvuglia et al. 2020 | Adaptation under a no mitigation policy | Nature-based solution tested under RCP8.5 | Baseline 'no heatwave' scenario | Consulted municipal policymakers for feedback after model completion |
|  |  |  | 30% housing with green roofs |  |
|  |  |  | 60% housing with green roofs |  |
|  |  |  | 100% housing with green roofs |  |
|  |  |  | Each scenario above tested under 1.5°C or 3°C building internal temperature reduction |  |
| Mixed policies |  |  |  |  |
| Dimitrova et al. 2022 | Mitigation policy | Climate target, emissions reductions, and behavioural change | National mitigation policy until 2030, current air pollution policies, and no additional support for clean cooking fuels | N/A |
|  |  |  | National mitigation policy until 2020 then mitigation measures to keep temperatures <2°C, current air pollution policies, and no additional support for clean cooking fuels |  |
|  |  |  | As above but with support for 15% LPG* stoves and 75% LPG fuel subsidies |  |
|  |  |  | National mitigation to 2020 with additional air pollution controls for industrial, power, household, and agriculture sectors (Maximum Feasible Reduction, MFR) but no support for clean cooking fuel transition |  |
|  |  |  | As above but with support for 15% LPG stoves and 75% LPG fuel subsidies |  |
| Scenario-based policies |  |  |  |  |
| Stephen & Barnett 2017 | No mitigation policy | Climate change emissions scenario | Baseline conditions | N/A |
|  |  |  | A1F1 |  |
| Weyant et al. 2018 | Mitigation policy | Climate change emissions scenario | Baseline - public health measures | N/A |
|  |  |  | RCP8.5 |  |
|  |  |  | Maintain global mean temperatures at <2°C |  |
| Pimpin et al. 2018 | Mitigation policy | Emissions reduction scenarios | Baseline - no change scenario | Government advisors on project steering committee and expert-led advice on data |
|  |  |  | Individual's PM<sub>2.5</sub> and NO<sub>2</sub> exposure reduced to natural (non-anthropogenic) levels |  |
|  |  |  | Individual PM<sub>2.5</sub> and NO<sub>2</sub> concentrations are reduced by 1 µg/m<sup>3</sup> |  |
|  |  |  | Individual NO<sub>2</sub> exposure reduced to EU limit of 40 µg/m<sup>3</sup> |  |
| Symonds et al. 2019 | Mitigation policy | Emissions reduction scenarios | Complete elimination of anthropogenic PM<sub>2.5</sub> | Expert-led advice on data |
|  |  |  | Compliance with WHO** guidelines for the annual mean PM<sub>2.5</sub> to not exceed 10 µg/m<sup>3</sup> |  |
|  |  |  | UK emission reduction in line with EU for baseline reduction of 3.6 µg/m<sup>3</sup> |  |
| Zeng et al. 2022 | No mitigation policy | Climate change emissions scenario | Baseline - no change scenario<br>RCP8.5 | N/A |

\*Liquid petroleum gas
\*\*World Health Organisation

Most projections were simulated under scenarios that included mitigation policies (n=4), although three models evaluated outcomes under a ‘no-mitigation policy’ future scenario. Only one model simulated adaptation action in the form of stepwise applications of a nature-based solution under the RCP8.5 scenario (Marvuglia et al. 2020). Most mitigation policies were applied homogenously to all synthetic individuals, apart from one study in which the simulation of a behavioural shift to cleaner cooking fuels was based on household income and maternal education level of individuals (Dimitrova et al. 2022) and a second study in which a building intervention, green roofing, determined personal temperature exposure (Marvuglia et al. 2020). A dynamic model implemented a time lag for air pollution reduction effects spanning the 20-year simulation, with its greatest effects reported 2-5 years after policy implementation (Symonds et al. 2019).

### Stakeholder engagement & economic evaluation

Three studies relied on expert or stakeholder engagement to inform model development and policy design (Table 3). Authors from two separate models on air pollution impacts each consulted with experts from the UK Committee on the Medical Effects of Air Pollutants (Pimpin et al. 2018, Symonds et al. 2019) and received guidance on model development from public health authorities through a project steering committee (Pimpin et al. 2018). Policymakers from four participating municipalities were convened to feedback on the simulation results of a multi-city research project after the research was completed (Marvuglia et al. 2020).

Two studies employed economic and costing analysis (Table 4). Health costing was applied to forecasts of the disease burden under a given climate scenario by evaluating QALYs lost, economic value of years of life lost (YLL), and healthcare costs (Stephen & Barnett 2017). Only one study applied cost-consequence analysis (CCA) to support the evaluation of alternative policy strategies (Pimpin et al. 2018). Both studies applied a social discount rate to weight future costs, at 3% and 1.5% respectively.

**Table 4.**
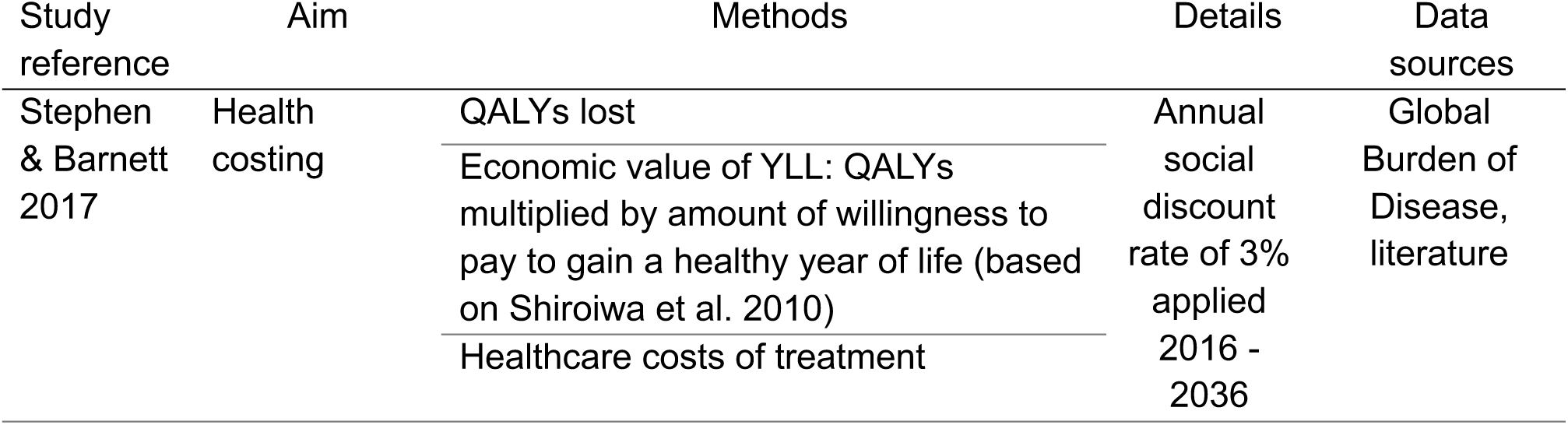
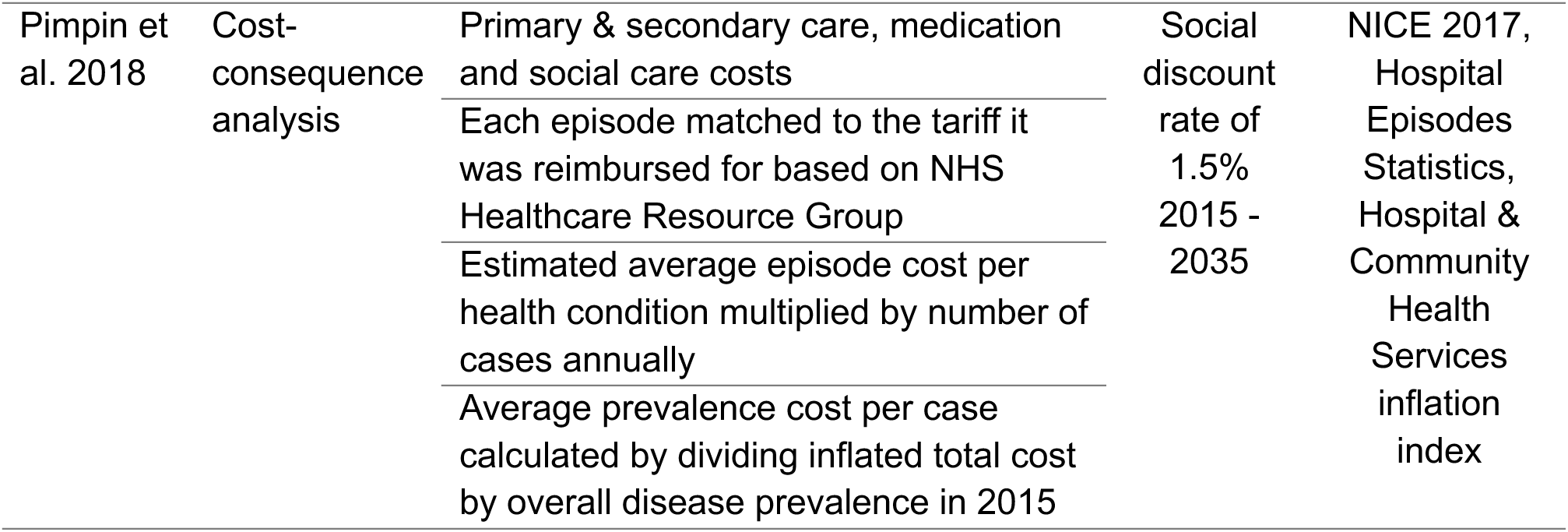
Economic and costing methods and data sources.

### Uncertainty analysis & validation

All models acknowledged uncertainty of parameters, and five studies reported methods and results of analyses conducted (Stephen & Barnett 2017; Zeng et al. 2022; Weyant et al. 2018; Pimpin et al. 2018; Dimitrova et al. 2022). Uncertainty was communicated through confidence intervals, credible intervals (for incidence), or uncertainty intervals and one model reported the min-max range for parameters (Marvuglia et al 2020). Two studies reported the use of Monte Carlo methods for sensitivity analysis (SA; Weyant et al. 2018; Zeng et al. 2022). Two models did not report uncertainty analysis (Symonds et al. 2019, Marvuglia et al. 2020), however Symonds et al. (2019) addressed this through a discussion of the extensive power needed to conduct sample-based SA for their dynamic model, with its large synthetic population and long projection period. Several studies confirmed that uncertainty in all input parameters was not captured in the uncertainty analysis (Dimitrova et al. 2022; Pimpin et al 2018; Stephen & Barnett 2017), however two studies reported comprehensive one-way SA on input parameters.

All studies bar one reported some type of validation of model components relating to either population synthesis or health outcomes. Most validation was conducted on health components and compared to estimated values from the literature or expert group consultations.

### Critical appraisal

All seven studies underwent critical appraisal as a component of the systematic review. Reviewer scores for most of the assessed criteria were high, indicating confidence in indicator criteria being met (Figure 2). Five indicators received low scores from multiple studies: methods of population synthesis reporting, validation reporting, reporting limitations of policy simulations, realistic achievability of (simulated) policies, and reproducibility. Certain criteria were commonly met among the study set, these included: acknowledgment of model limitations, reporting a counterfactual simulation (or baseline conditions), justification of source of exposure-response association data, description of uncertainty analysis, citing data sources for population synthesis construction, and reporting key model assumptions.

**Figure 2.**
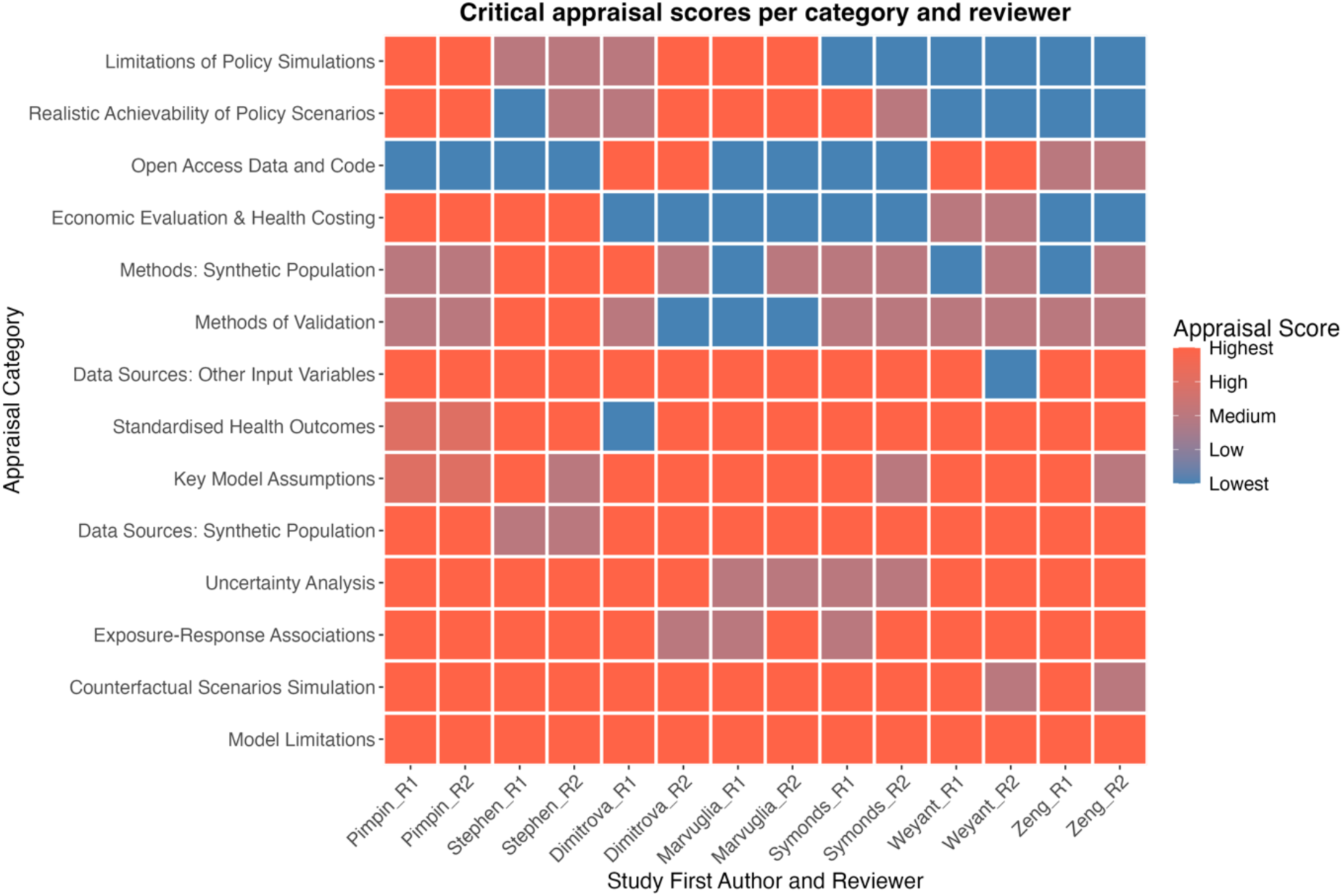
Heatmap of critical appraisal scores for the included set of studies.

## Discussion

The evolving use of spatial MSM methods for data-driven climate-health impact assessment and policy evaluation motivated this review of its applications and despite a limited number of published studies, the consistency of methods suggests a shared framework. In this discussion, we position these MSM models in the context of current critical issues in climate-health impact assessments and climate policy evaluation, following the three review objectives. For climate modellers and policy actors considering the value of MSM as a decision-making tool, we provide a discussion of its limitations and future opportunities.

### Model characteristics

A defining characteristic, and caveat, of MSM models is their relative complexity. To plausibly inform policy decisions, these methods depend on identifying multiple representative data sets that are sufficiently disaggregated to inform model parameters at the level of an individual. In doing this, a balance must be found between incorporating enough detail to achieve credible outcomes and avoiding over-parameterisation, which can complicate model calibration and reduce generalisability. To that end, decisions related to model set up and the assumptions entailed are important considerations for prospective modellers and policymakers, and in this review, some trends stood out.

In general, population cohorts were more commonly used than whole society simulations; sometimes this was done to focus on a particularly vulnerable group, but in doing so a key asset of MSM models was underutilised, that is their ability to illustrate distributional impacts across a population. Dynamic methods were more frequent, allowing the lifecycle of individuals to be tracked through the simulation, however most dynamic models used annual time-steps which limited shorter-term assessments of risk and potentially underestimated certain outcomes. Single climate hazard assessments were common, ignoring the complexity of interdependencies, and delicate balance, among Earth’s systems. Some models averaged exposures over wide geographic areas, such as whole municipalities, to make up for missing high-resolution climate data; mixed climate policies were rarely tested in favour of single scenarios or interventions. These choices were influenced by data availability, model run times, and gaps in understanding of interactions or relationships between variables.

The decision to utilise data intensive methods like MSM rests on its ability to capture the complexity and diversity of individual experience and to demonstrate how this experience impacts population estimates, necessarily restricting their use to contexts where highly detailed data and infrastructural capacity is available. That said, for policymakers most concerned about existing population disparities and the equity impacts of climate policies, even when based on simplifying approaches, MSM models may still yield more relevant insights than the portrayal of homogenous risks and vulnerabilities otherwise would. These insights may provide a more realistic basis for climate decision-making, potentially improve governance, and can also highlight where pressing data gaps hamper evidence-informed policymaking. In this sense, climate modellers and policymakers considering if MSM methods are appropriate for their needs should consider their resources, but also if their primary objectives warrant this micro-level approach.

### Exposure-response relationships

Exposure-response curves are integral to environmental risk assessments as they quantify the average expected response per unit of an observed exposure. In climate-health impact assessments, the health response is conditional on the distribution of covariates in the exposed population that the data was collected from, and these may differ between populations. In addition, exposure-response curves represent population-averaged values of response; when applied to individual-level simulations, these functions can obfuscate the individual variability in response – precisely the type of detail MSM methods are designed to uncover.

In settings with limited data availability, exposure-response relationships drawn from meta-analyses of global literature reviews have facilitated climate-health impact assessment where it may not have been previously possible. Though the use of data from mixed populations with distinctive covariate distributions raises some questions of the accuracy of estimated health response associations, this may be justifiable in situations where data scarcity may otherwise impede progress.

### Uncertainty

Reporting uncertainties strengthens confidence in policy analyses. Understanding the limits of imperfect knowledge can guide decision-makers as to whether additional resources, such as investments in data collection or knowledge support, are needed to strengthen model forecasts (Pianosi et al. 2016). Our review indicated that policy simulation results were typically accompanied by quantitative uncertainty analyses (UA) or at least a discussion of sources of uncertainty; however, difficulties in implementing standard approaches to UA due to long run times and processing power needs were also reported. Since reliability of modelled outputs underpins the evidence-to-policy pathway, insufficient exploration of uncertainty could compromise its value for decision-support. Where quantification of uncertainty is limited, discussion of these limitations and the resources needed to address them should be prioritised.

### Scope of climate policies

Given the history of MSM for policy testing, the eligibility criteria of this review required evidence of policy evaluation for inclusion. Reflecting a previously reported limitation of climate modelling studies (Whitmee et al. 2024), most models in our study set applied a single mitigation action and only one study tested an adaptation action. Among mitigation studies, only one employed a mixed policy approach, applying multiple combinations of mitigation actions including a national carbon tax, policies to induce behavioral change in the cooking fuel transition, and structural end-of-pipe emissions reductions in the form of MFR policies (Dimitrova et al. 2022). Two earlier MSM studies also simulated health co-benefits of air pollution reduction targets and discussed synergies between air quality improvements and low carbon climate policies but did not attempt to estimate the impacts of GHG emission reductions on co-emitted pollutants and subsequent feedback loops. While relationships between climate change and air pollution are complex, evidence suggests that reaching the recommended annual air quality exposure target of 5μg/m^3^ (WHO 2021) will not be possible for some highly industrialized countries using end-of-pipe controls alone and climate mitigating clean energy policies will invariably be necessary (West et al 2013; Xing et al. 2020). A recent review of climate policy effectiveness to reduce GHG emissions found that ‘policy mixes’ generally had greater effect sizes than standalone policies, further underscoring the importance of integrating combinations of climate policies into simulation models to remain relevant for decision-making (Stechemesser et al. 2024).

The diversity of mitigation actions also presents challenges for comparative evaluation between models. To address this, a typology of climate policies has recently been elaborated categorizing actions into four groups: specific interventions; unspecified actions that are assumed to induce a structural change; those that induce a behavioral change; or unspecified actions that realize a particular climate target (Reynolds et al. 2025). In our study set, dominance of the latter policy actions underscores that the selected studies may be more useful for advocacy on the urgency of the climate threat than as practical decision-support tools for specific interventions.

This distinction is critically important as the world remains offtrack to meet Paris Agreement goals, with little evidence available to demonstrate specific health gains in vulnerable populations that could strengthen the argument for policymakers wishing to act on GHG emissions. There is considerable opportunity to provide more realistic climate policy evaluations using MSM methods as their construction permits multiple policies to be implemented at different model stages and integration of outputs from other models. Presently, this potential may be limited by gaps in understanding of interactions between and compounding effects among different policy actions; the latter is a known cause of underestimated outcomes and frequently unaccounted for in mixed sector policy evaluations (Stechemesser et al. 2024). Dynamic designs also offer specific opportunities to enhance policy simulations less easily afforded to static models: their ability to temporally incorporate lagged effects and feedback loops. The former, illustrated as delayed health impacts of air pollution reduction policies by Symonds et al., serves as an example of more realistic policy scenario building (2019).

Notably, every model in our SR that incorporated climate emissions scenarios relied on projections under RCP8.5 and its predecessor A1F1, and just one model compared the resulting health impacts to those under a low-emissions scenario in which global mean temperatures <2°C are maintained (Weyant et al. 2018). Among RCP scenarios, the ‘high-risk future’ denoted by RCP8.5 corresponds to the 90^th^ percentile of baseline forcing levels and represents a world of intensive fossil fuel reliance with no mitigation efforts (van Vuuren et al 2011; Rogelj et al. 2012). The choice to use this scenario raises questions about the credibility of policy guidance in simulations that are anchored in a low-probability future (Hausfather & Peters 2020); with real-world examples not available to draw on, simulations conducted under multiple climate scenarios, and exploring a range of possibilities, would improve the policy plausibility of modelled outcomes.

### Stakeholder engagement

Technical and scientific outputs are generally strengthened by integrating relevant stakeholders and experts into collaborative methods for research design (co-design), data collection and results analysis (co-production), and dissemination of outputs (co-dissemination) (Gerlak et al. 2023). These are important components of procedural justice, which aims to improve fairness, accountability and inclusivity (Newell et al. 2021). Investigating the health effects of climate change is a complex process and some aspects can be contentious - engaging a diverse collection of stakeholders lends legitimacy to research where gaps in knowledge may otherwise hinder progress, improves buy-in from affected communities, and can lead to more salient and context-specific findings (Meadow et al. 2015). In LMICs, where the trade-offs of budget deployment decisions may compromise programs elsewhere, this engagement is critically important.

Across the selected studies, we found a moderate level of stakeholder engagement, primarily with academic and government-based experts, occurring in different phases of the research process. Authors that sought feedback from municipal decision-makers on their MSM modelling of green roofing to reduce heat mortality reported that few were convinced of its suitability for implementation, potentially implying that engagement earlier on at the co-design stage of MSM development may have been desirable. Conspicuously, models of LMIC populations rarely reported any stakeholder engagement methods; greater support may be needed to facilitate these collaborations.

### Economic and cost analysis

To enhance strategic decision-making, the opportunity costs of budget allocation choices should be reported using established methods of economic evaluation. Given the long history of MSM applications in tax-benefit modelling, the scarcity of applied economic evaluation methods in our study set was unexpected. Only one study compared costs and outcomes to distinguish between alternative policy options and not coincidentally given policymakers attention to cost implications, this study alone engaged public health civil servants on a steering committee for its model development (Pimpin et al. 2018). Discounting, important for estimating the present value of future health and healthcare costs, was applied variably in two studies examining either healthcare costs alone or cost and health outcomes. Notably, a third study described the use of discounting for health outcomes projections in its supplementary materials only (Weyant et al 2018) with no accompanying cost analysis or explanation. The variation between these examples highlights that economic methods have not been properly considered or fully integrated into these studies. There are valid reasons for its limited use in climate-health impact assessments, primarily, the lack of consensus on how to value health gains of climate action (Scovronick et al. 2020; Cromar et al. 2021), and a lack of best practice for effective economic frameworks for climate policy evaluations (Cubi-Molla et al. 2025). Either of these may deter authors from undertaking comprehensive economic evaluation, however as our review findings suggest, involving decision-makers and policy actors early in the model development process may still improve its economic utility overall in decision-support contexts.

### Challenges and future directions

The opportunity to combine dynamic and spatial methods within an individual-based model offers a promising method to capture temporal, geographical, and personal heterogeneity. It also provides an approach to policy simulation that can integrate considerations of distributive and procedural justice and trade-offs against competing demands for scarce resources. This specificity, however, comes with steep technical, data, and infrastructural costs, challenging its widespread use. Some opportunities to facilitate their methodological advancement and mainstreaming warrant reflection.

#### 1. Accessibility of methods

Our review confirms that these methods appear to be more commonly applied to high-income populations likely reflecting the high existing technical capacity and support for evidence-to-policy collaborations, greater accessibility to detailed open-access spatial microdata sets, and availability of HPC facilities for research use. The costs of such capacity needs are enormous, with a recent review citing multimillion dollar budgets for the creation of some governmental dynamic MSM models in the USA (Harding 2023). This leaves these methods largely out of reach for modellers and decision-makers in low resource countries, which bear a disproportionate vulnerability to climate change and where effective climate policies are most urgently needed (Naser et al. 2024; Ebi & Luchters 2021). Dedicated efforts to streamline these methods and greater support for international research collaborations, as well as expanded funding that prioritises policy simulation research, could begin to address challenges of accessibility.

#### 2. Exposure-response relationships

Exposure-response relationships typically represent the average population response per unit of exposure. When applied at an individual level, some individual variation is obscured. To explore the effect of covariate distributions, growing interest has focused on individual conditional expectation (ICE) curves (Goldstein et al. 2015, Molnar 2025). These visualise covariate heterogeneity by plotting the response curves per individual to show the range of expected response probabilities per unit of exposure, conditional on selected covariates. These methods may represent an opportunity to improve corresponding impact estimates by accounting for some individual heterogeneity in exposure-response distributions.

#### 3. Reproducibility

Critical appraisal of models included in our review identified limited reporting for reproducibility. However, sharing code and datasets improves transparency and could support the growth and development of the climate-health MSM discipline by permitting successive model iterations to be built upon. Sharing code for complex models additionally lowers barriers for researchers based in low-resource contexts.

#### 4. Modelling for policy cycles

The lengthy timeframes for research dedicated to complex public health issues sit sharply in contrast to short timelines of policy cycles (Whitty 2015, Berry et al. 2018). This misalignment may be an obstacle for the use of data-intensive methods like MSM for policy development, despite their academic appeal. However, decision-makers often do not need the same degree of technical detail and validation to guide funding allocations and therefore modellers may consider employing more efficient methods of construction. Since MSM has been used in policy modelling for some time, expanding or building on existing components such as attribute-rich synthetic populations or integrating sub-models to generate specific exposure estimates could speed development of new climate-health applications. Given the importance of evaluating opportunity costs when comparing policy options, embedding economic evaluation into these models as a standard practice would also improve the decision-making process. Toward this objective, standards for economic evaluation of health co-benefits of climate change are currently being developed through a global consensus initiative (ECO-CHICA, 2025).

#### 5. National risk assessments

National climate risk assessments have gained increasing attention from intergovernmental organisations focused on improving population health, poverty alleviation and sustainable development (WHO 2021, Zebish et al. 2023). These assessments encourage the use of quantitative methods to measure health risks and vulnerabilities to strengthen the evidence for national and sub-national policy prioritisation efforts and are particularly important in LMICs. Individual-based methods could be used to compare and contrast health burden impacts associated with different climate hazards under future climate scenarios to facilitate more informed resource allocations and improve equity considerations in climate policy decision-making and financing negotiations.

### Limitations of this review

While the use of systematic review methods aims to reduce bias and produce a comprehensive assessment of the literature, some limitations should be noted. First, MSM models largely remain within the domain of government policy meaning that our review of published and peer-reviewed papers may be biased towards academic outputs. However, academic research is less constrained by budget priorities and can be more agile and open to collaborative ideas at the forefront of methodological advances, therefore the likelihood that this bias resulting in ‘missed’ examples is small. Our focus on peer-reviewed publications also raises the quality and consistency of methods within the study set, which is necessary for meaningful comparison. Second, spatial methods were consistently applied and reported among studies published from 2018 onwards, prompting a protocol amendment for an earlier study that otherwise met eligibility criteria. This might suggest that as these methods gain wider acceptance, more uniformity of methods will occur. Third, as our eligibility criteria required inclusion of a climate policy, this necessarily excluded some MSM models that strictly assessed health impacts of climate hazards. This criterion was applied in recognition of the history of MSM for policy decision-support as well as the pragmatic importance of identifying actions that can be implemented today to avoid further carbon lock-in and to reduce future health consequences. As these methods are a relatively recent addition to the climate-health modelling arsenal, it remains clear that some further refinement may be necessary with their continued development.

### Conclusions

We set out to characterize the evolving use of MSM methods as a tool to quantify health impacts of exposure to climate hazards and to provide decision-support for climate policies. Evidently, the promise of this methodology is currently constrained by limited scholarship, although a common framework is emerging. Important challenges relate to accessibility of the methods, credibility of climate policy simulations, and model reproducibility, requiring some further development. It is apparent that the global response to climate change requires more action and greater collaboration than ever before; to this end, MSM methods offer to enrich the science-policy space and improve the visibility of vulnerable groups in climate policy modelling. Decision-makers and stakeholders working on climate policies and concerned with justice might consider the use of MSM methods to strengthen the evidence describing health impacts of climate change and to provide a quantitative basis to support resource allocation decisions.

## Data availability statement

All data to support the methods and findings of this study are available as supplementary materials.

## Acknowledgement

No specific grant from funding agencies in the public, commercial, or not-for-profit sectors was received for this work.

## CRediT statement

Ariel Brunn: Conceptualisation, Methodology, Investigation, Data Curation, Formal Analysis, Visualisation, Writing - Original Draft, Writing - Review and Editing; Roberto Picetti: Methodology, Investigation, Data Curation, Writing - Review and Editing; Lauren Ferguson: Methodology, Investigation, Data Curation, Writing - Review and Editing; Francis Ruiz: Methodology, Writing - Review and Editing; Petra Meier: Methodology, Writing - Review and Editing; Rosemary Green: Methodology, Writing - Review and Editing; James Milner: Methodology, Writing - Review and Editing. All authors have read and agreed to the published version of the manuscript.

