## Supplementary materials for "Estimating the health impacts of climate change for policy decision-support: a systematic review of spatial microsimulation methods"

Table S1. Climate and health data sources, metrics and exposure-response relationships

| Study reference | Climate hazard | Data source | Health outcome | Health metric | Data source | Exposure-response relationship | Method or source |
|---|---|---|---|---|---|---|---|
| **Stephen DM 2017** | Daily minimum and maximum ambient temperature & daily rainfall | AU Bureau of Meteorology, projections from CMIP3 (A1F1) | Salmonellosis morbidity & sequelae (post-infectious IBS and reactive arthritis) | QALYs, case incidence | Queensland Health | Temp/rainfall combinations and cases | Poisson regression with lags |
| **Weyant et al 2018** | Atmospheric $CO_2$-induced iron and zinc losses in food crops | FAOSTAT, USDA, IPCC ($CO_2$) | Iron deficiency anemia, zinc deficiency-related diseases: pneumonia, malaria, lower respiratory infections | YLDs, YLLs, DALYs | Global Burden of Disease study (GBD) | Nutrient intake and health outcomes | GBD |
| **Pimpin et al 2018** | Annual averaged $PM_{2.5}$ & $NO_2$ | Land-use regression models (de | Ischemic heart disease (IHD), stroke, lung | Incident cases per 100,000 | UK Office of National Statistics, Public | $PM_{2.5}$ and health outcomes | Literature, expert group |

| | | | | | | | |
|---|---|---|---|---|---|---|---|
| | | Hoogh K *et al*. 2016 & Gulliver J *et al*. 2013) | cancer, COPD[†], dementia, diabetes mellitus (type II), asthma, low birthweight | | Health England, Alzheimer's Society Dementia UK, British Heart Foundation, British Lung Foundation, Cancer Research UK | | |
| **Symonds et al 2019** | Annual averaged $PM_{2.5}$ | ADMS pollution dispersion model (Ricardo Energy & Environment 2017) | IHD | Mortality per 100,000 | GBD | $PM_{2.5}$ and IHD | Literature, expert group |
| **Marvuglia et al 2020** | Apparent temperature estimated from daily ambient temperature and monthly dewpoint | Urban Atlas (EU Copernicus Programme); Projections from CMIP5 (RCP8.5) | Heat-attributable mortality | Annual cumulative mortality, percent mortality | Estimated using formulas from Baccini M *et al*. 2008 | Temperature & humidity via a new estimated variable, the apparent temperature, and heat mortality | Literature |

| | | | | | | | |
|---|---|---|---|---|---|---|---|
| **Zeng et al 2022** | Ambient temperature-influenced food crop availability | Results from IMPACT 3.3 (RCP8.5); China Health and Nutrition Survey (2004 – 2011) | Gastric, lung and esophageal cancer, change in body mass index (BMI) | DALYs per 1000 | Global meta-analyses (GBD) | Dietary intake and health outcomes | GBD |
| **Dimitrova et al 2022** | Annual averaged $PM_{2.5}$ and household air pollution | $PM_{2.5}$ (Hammer et al. 2020) and MESSAGE-GLOBIUM GAINS module for projections; cooking fuel type from India National Family Health Survey | Stunting via in-utero exposure | Cumulative preventable number of stunted children; Percent change in stunting prevalence | India 2015 - 16 Demographic Health Survey (NFHS) | $PM_{2.5}$ and stunting | Logistic regression |

### S2. Critical Appraisal Key

***Objective:*** *To evaluate the validity of model methods and sources of data to provide an accurate and reliable assessment of health impacts associated with climate exposures at an individual level and to assess the overall credibility of the model to provide a realistic representation of climate change policy impacts.*

***Article Identifier***
First author and year of publication

***Validity of model parameters***
1. Are key assumptions described?
    - Are the assumptions justified?
    - Score = 0.5 if assumptions are listed but not substantively justified
    - Score = 1 if justified/discussed by authors

2. Are data sources for synthetic population construction cited?
    - Are data sources for the synthetic population cited?
    - Score = 1 if cited

3. Are the methods of population synthesis reported?
    - Are recognised methods of population synthesis reported[1]?
    - Score = 0.5 if any methods of population synthesis are reported
    - Score = 1 if recognised methods are reported

4. Have data sources for input variables (health, socioeconomic, climate exposures) been listed?
    - Have data sources been cited?
    - Score = 0.5 if only one cited
    - Score = 1 if all are cited

5. Have health metrics been standardised?
    - Are the health metrics justified and standardised (e.g. rate per 100,000, where appropriate)?
    - Score = 0.5 if justified
    - Score = 1 if health metrics are standardised

6. Was the source of exposure-response relationships justified or explained?
    - Example discussion points:
        - The associations were obtained from meta-analyses or estimated from empirical data sources (versus taken from literature)
        - Associations were based on local data and appropriate for the context of the simulation

[1] Methods described in Lomax N (2022) Microsimulation, Handbook of Spatial Analysis in the Social Sciences. In: Rey S & Franklin R (eds), Edward Elgar.
doi: https://doi.org/10.4337/9781789903942

    - The authors included descriptions of how exposure-response relationships vary among groups (e.g. age-related), if relevant
  - Score = 0.5 if secondary data used (from literature)
  - Score = 1 if associations were generated from empirical data or taken from meta-analyses, or if substantive discussion and validation justifies use of associations from the literature

7. Has any evaluation of uncertainty in parameters or model methods been conducted?
   - Have sources of uncertainty in the parameters themselves, or in the model methods, been considered?
   - Has any uncertainty analysis been conducted to demonstrate the range of potential outputs of the model (at some probability level)?
   - Has the impact of uncertainty on model outputs been considered through sensitivity analysis?
   - Score = 0.5 if discussion of sources of uncertainty in parameters and/or model methods is present
   - Score = 1 if methods have been undertaken to demonstrate uncertainty, such as sensitivity analysis

8. Have methods of validation been conducted and reported?
   - Has the constructed synthetic population been validated?
   - Have health outcomes been validated?
   - Score = 0.5 at least one component (population or health outcomes) validated
   - Score = 1 all components validated

9. Are limitations of the model described?
   - Score = 1 if present

***<u>Credibility of climate policy simulation</u>***

1. Are counterfactual scenarios simulated?
   - Score = 0.5 if counterfactual scenarios were implemented
   - Score = 1 if counterfactual scenarios were implemented considering demographic change

2. Is there an assessment or discussion of the realistic achievability of the scenarios / pathway to reach change / potential scale-up?
   - Example assessment points:
     - Has the policy already been implemented?
     - Have stakeholders or policymakers been consulted or co-produced the policies?
     - Are incremental policy options simulated (up to a pre-identified threshold)? As an example, a policy might simulate 10%, 15%, or 20% uptake within the population.
     - Is the policy implemented for particular vulnerable groups or is a blanket population approach used for the simulation (if the latter, is an all or none approach justified)?

- Are the baseline timeframe and projected horizon justified by authors?
    - Are equity impacts of policy uptake considered?
    - Are 'real-world' issues associated with policy implementation discussed, referring to empirical data where possible?
- Score = 0.5 if only one point is implemented by authors
- Score = 1 if at least two of the above points is implemented by authors

3. For discounted valuation estimates, justification of choice of fixed or variable rates and sources described for specific rates chosen
    - Is sensitivity analysis conducted for discounted rates, including at least rates of 0% and 3%?
    - Score = 0.5 if present
    - Score = 1 if justification is available or if sensitivity analysis conducted

4. Are limitations of the policy simulations discussed?
    - Score = 0.5 if acknowledged
    - Score = 1 if discussed substantively

***Reproducibility***

When legally and ethically possible, openly and publicly share data and code used. If there are limitations to data sharing, have these been stated?

- Score = 0.5 if limitations on data sharing is stated
- Score = 1 if data and code is shared

***Critical appraisal score***

Full score = 14
12 - 14: high confidence in model and/or policy validity and credibility
10 - 11: moderate confidence in model and/or policy validity and credibility
0 - 9: low confidence in model and/or policy validity and credibility

***References***

Picetti R, Juel R, Milner J, Bonell A, Karakas F, Dangour AD, Yeung S, Wilkinson P, Hughes R. Effects on child and adolescent health of climate change mitigation policies: A systematic review of modelling studies. *Environ Res*. 2023 Dec 1;238(Pt 1):117102. https://doi.org/10.1016/j.envres.2023.117102.

Hess JJ, Ranadive N, Boyer C, et al. Guidelines for Modeling and Reporting Health Effects of Climate Change Mitigation Actions. *Environ Health Perspect*. 2020;128(11):115001. https://doi.org/10.1289/EHP6745

Lomax N (2022) Microsimulation, Handbook of Spatial Analysis in the Social Sciences. In: Rey S & Franklin R (eds), Edward Elgar. doi: https://doi.org/10.4337/9781789903942

### S3. Protocol for a systematic review of spatial microsimulation modelling applied to health impact assessment of climate change policies

*Background*

Microsimulation modelling (MSM) is an individual-based method frequently used to guide health policy decision-making. It is particularly useful in the assessment of distributional impacts of policy applications, facilitating comparative evaluation without the need for lengthy or costly field trials. Recently, an emerging area of MSM has focused on estimating the impacts on health outcomes arising from climate change mitigation and adaptation actions.

Several features of MSM make it a natural companion for climate policy evaluation. For instance, the fine scale of spatial MSM is conducive to matching geographic variations in climate with geo-located individuals, capturing the range of personal exposures experienced at local levels. In addition, the process of simulating individuals through the creation of a 'close-to-reality' synthetic population improves the granularity of social attributes that drive health vulnerability in specific sub-populations, better reflecting the inequities important to climate justice debate (Thomas et al 2018; Kopasker et al 2023). Together, these attributes yield highly specific aggregated population estimates of disease burdens attributable to climate exposures.

However, for the climate policy community, the centrality of policy testing to MSM is its most compelling feature. MSM has been used for decades as a policy testing tool allowing decision-makers to systematically and incrementally compare and contrast interventions to optimise the policy development and design process (Rutter et al 2011; Schofield et al 2018). For policymakers working on climate mitigation and adaptation, MSM may be used to explore promising sector-specific policies and the range of their impacts among vulnerable demographics, as well as potential trade-offs inherent to optimising health and environmental outcomes amid limited budgetary allocations.

With foundering progress in implementation of climate policies and current targets for greenhouse gas emissions insufficient to meet Paris Agreement temperature aims, attention is turning to the policymaking process itself and efforts to improve its effectiveness and scope (OECD 2024). The emerging use of spatial MSM as a tool for data-driven climate policy scenario testing warrants a review of its applications, methodological trends, and, critically for decision-makers, it's limitations.  This systematic review aims to summarise existing spatial MSM models[2] in the peer-reviewed literature and to draw conclusions intended to guide progression of this evolving area of policy simulation modelling.

[2] This review will focus on spatial microsimulation models, defined by Lovelace & Dumont (2016) as "the creation, analysis and modelling of individual level data allocated to geographic zones" and will focus on models incorporating a synthetic population and simulations of interactions between individuals and their spatial environment. See expanded model inclusion definition on page 2.

*Objectives*
The specific objectives of this SR of the peer-reviewed literature are:
1) To describe characteristics of MSM that evaluate the health impacts of simulated climate change policies, including methods used for synthetic population generation and exposure-health associations.
2) To assess the scope of climate change policies applied in the included study set of peer-reviewed literature.
3) To appraise the validity, scope and current challenges and to make recommendations to guide future model development for health impact assessment of climate change policies using MSM methods.

*Protocol*
An *a priori* protocol was developed using the Preferred Reporting Items for Systematic Reviews and Meta-Analyses for Protocols (PRISMA-P; Shamseer et al. 2015) to reduce sources of bias in the review. The search strategy included three subjects, namely, MSM methods, health outcomes, and climate change policy.

| Subject | Term | Synonyms |
|---|---|---|
| Methodology | microsimulation | "synthetic population" OR "individual-based" |
| Outcome | health | disease OR wellbeing OR burden |
| Policy | climate change | environment OR weather OR "global warming" OR "climate crises" |

*Search Terms*
The search string was formulated through an iterative process in two bibliographic databases, MEDLINE and Web of Science to identify a list of known publications that matched the inclusion criteria and is detailed in Table 1A (Appendix).

Five bibliographic databases, MEDLINE, Embase, EconLit, Scopus and Web of Science, will be searched for peer-reviewed published literature. Google Scholar and the International Journal of Microsimulation will also be searched to identify additional peer-reviewed reports. Grey literature will not be included as an objective of this review is to assess the scope and breadth of policy testing in MSM present in the academic literature and to comment on its potential as a tool for decision-makers to drive and inform credible policy development.

Abstracts in languages other than English will not be included; recent literature reviews have found that MSM research intensity is overwhelmingly based in the USA, United Kingdom and Australia suggesting that exclusion of non-English language articles is unlikely to be a serious constraint (Schofield et al 2018; Mertens et al 2022; Oliveira et al 2024). Studies published in any location or featuring a synthetic population from any region worldwide will be included, however only those published over the past decade will be reviewed. This timeframe corresponds to the historic adoption of the Paris Agreement in 2015 by 196 Parties and concomitant requirements for countries to commit to climate action planning via their Nationally Determined Contributions (NDC).

Only modelling studies will be included which specify the use of spatial MSM methods or that are identifiable through the inclusion of simulations involving a synthetic population and that describe spatial interactions directly between an individual and its environment. In this review, agent-based models (ABM) will not be included and although some degree of taxonomic ambiguity exists between ABM and MSM, the former are identified as individual-based models which simulate interactions among individuals and may involve smaller sub-populations due to their increased complexity and computational power needs (Lovelace & Dumont 2016).

*Data Selection and Management*
Reference management and screening will use Rayyan software for de-duplication and screening of articles. Title screening will be conducted by a single independent reviewer to screen out excluded study designs if listed in the title, e.g. "Systematic Review". Abstract screening will be conducted by two independent reviewers to identify studies that meet criteria for full-text review. Positive response to three abstract screening questions will advance a study to the next stage of screening. Full-text review will be undertaken by two independent reviewers, who will also document reasons for study exclusion.

*Inclusion Criteria*

| **Criteria** | **Inclusion Criteria** |
|---|---|
| Publication Year | 2015 - 2025 |
| Location of Study | Any location accepted |
| Language | English only |
| Study type | Spatial MSM; all other study types excluded. |
| Publication type | Peer-reviewed literature only; full-text must be available |
| Population | Any synthetic individual-based population |
| Exposure | Any climate variable such as ambient temperature, precipitation, particulate matter etc. |
| Comparator | Counterfactual climate scenario; studies that do not have this may still be included (for instance *ex-post* policy simulations) |
| Outcomes | Human health outcomes, expressed as a disease burden estimate, *e.g.* mortality or morbidity (DALYs, QALYs etc.), VSL |

*Abstract Screening Questions*
1) Does the article describe any individual-based model, such as microsimulation, agent-based model, or any simulation model utilizing a synthetic population?
2) Is the study about humans or human systems and environmental or climatic exposures (exclude if focus is on animal, insect, plant, genetic/cellular or transport systems)?
3) Is it a peer-reviewed journal article?

*Full-text Screening Questions*
1) Does the study describe a spatial microsimulation model (see definition, page 1)?
2) Is at least one of the modelled outcomes a human health outcome?
3) Does the model simulate an action, intervention, policy or strategy aimed at mitigating or adapting to climate change, or simulate projections using an established greenhouse gas emissions scenario?

*Data Extraction*

The data extraction template will be developed in Microsoft Excel. Three categories of data will be extracted, namely, 1) Article Meta-data: including title, citation, year of publication and region or country of simulated population; 2) Model Specifications: including type of climate exposure, health outcomes simulated, method of construction of synthetic population, model methods, exposure-response relationships and source, model projection time period, uncertainty analysis, and methods of validation, author-assessed limitations; 3) Policy Scenarios: type of and rationale given for simulated policies, net effects of modelled policies on baseline outcomes (i.e. whether policies positively or negatively impact outcomes) and a summary of any economic outcomes included.

*Critical Appraisal*

Included studies will undergo critical appraisal to evaluate evidence for model verification, management of uncertainty, internal and external validation, and credibility of simulated policy scenarios. The critical appraisal process will be conducted by duplicate reviewers using a custom-built tool, adapted from Picetti *et al*. (2023) and Hess *et al*. (2020).

*Synthesis of Results*

The screening process will be illustrated using a flow chart template. Meta-data will be charted to identify trends. Model methods will be tabulated (example Table 2A, Appendix). Climate change policies will be categorised by action (mitigation or adaptation) and simulation results will be summarised by impact (positive, negative, or no impact). If the model also simulated the impact of its policies on climate exposures in addition to health outcomes, this will be noted. The results of the critical appraisal process will be tabulated.

*Ethics and dissemination*

This review will use publicly available open-access reports and articles from bibliographic databases and online. As no primary or identifiable data will be used, ethical approval is not required. The findings of the review will be disseminated through publication online and in a peer reviewed journal such as Climate Policy, as well as at relevant meetings and workshops.

*Review Team*

Ariel A Brunn – Review Team Lead, LSHTM
Lauren Ferguson – Review Team, Screening, Harvard T.H. Chan School of Public Health
Roberto Picetti – Review Team, Data Extraction & Critical Appraisal, LSHTM

*Review Advisory Group*

Professor Rosemary Green, LSHTM
Professor Petra Meier, University of Glasgow
Associate Professor James Milner, LSHTM
Senior Policy Fellow Francis Ruiz, LSHTM

*References*

de Oliveira C, Matias MA, Jacobs R. Microsimulation Models on Mental Health: A Critical Review of the Literature. *Value Health*. 2024;27(2):226-246. doi:10.1016/j.jval.2023.10.015

Hess, J. J., Ranadive, N., Boyer, C., Aleksandrowicz, L., Anenberg, S. C., Aunan, K., Belesova, K., Bell, M. L., Bickersteth, S., Bowen, K., Burden, M., Campbell-Lendrum, D., Carlton, E., Cissé, G., Cohen, F., Dai, H., Dangour, A. D., Dasgupta, P., Frumkin, H., Gong, P., Ebi, K. L. (2020). Guidelines for Modeling and Reporting Health Effects of Climate Change Mitigation Actions. *Environmental health perspectives*, *128*(11), 115001. https://doi.org/10.1289/EHP6745

Kopasker, D et al. 2023. Microsimulation as a flexible tool to evaluate policies and their impact on socioeconomic inequalities in health. *The Lancet Regional Health – Europe*, Volume 34, 100758. https://doi.org/10.1016/j.lanepe.2023.100758

Lovelace, R, Dumont, M, 2016. Spatial microsimulation with R. CRC Press. https://spatial-microsim-book.robinlovelace.net/

Mertens E, Genbrugge E, Ocira J, Peñalvo JL. Microsimulation Modeling in Food Policy: A Scoping Review of Methodological Aspects. *Adv Nutr*. 2022 Mar;13(2):621-632. https://doi.org/10.1093/advances/nmab129.

OECD (2024), *The Climate Action Monitor 2024*, OECD Publishing, Paris, https://doi.org/10.1787/787786f6-en.

Picetti R, Juel R, Milner J, Bonell A, Karakas F, Dangour AD, Yeung S, Wilkinson P, Hughes R. Effects on child and adolescent health of climate change mitigation policies: A systematic review of modelling studies. Environ Res. 2023 Dec 1;238(Pt 1):117102. https://doi.org/10.1016/j.envres.2023.117102.

Rutter CM, Zaslavsky AM, Feuer EJ. Dynamic Microsimulation Models for Health Outcomes: A Review. *Medical Decision Making*. 2011;31(1):10-18. https://doi.org/10.1177/0272989X10369005

Shamseer L, Moher D, Clarke M, Ghersi D, Liberati A, Petticrew M, Shekelle P, Stewart LA, the PRISMA-P Group. Preferred Reporting Items for Systematic Review and Meta-Analysis Protocols (PRISMA-P) 2015: elaboration and explanation. BMJ 2015;349:g7647. doi: 10.1136/bmj.g7647

Schofield DJ, Zeppel MZB, Tan O, Lymer S, Cunich MM, Shrestha RN. 2018. A brief, global history of microsimulation models in health: Past applications, lessons learned and future directions. *International Journal of Microsimulation*. 11(1); 97-142. doi: 10.34196/ijm.00175

Thomas K, Hardy R D, Lazrus H, Mendez M, Orlove B, Rivera-Collazo I, Roberts J T, Rockman M, Warner B P, and Winthrop R. 2019 Explaining differential vulnerability to climate change: A social science review. *WIREs Clim Change*. **10** e565. https://doi.org/10.1002/wcc.565

**Appendix (Protocol)**

| | Database Search Results | | | | |
|---|---|---|---|---|---|
| **Search string** | **MEDLINE** | **Embase** | **EconLit** | **Web of Science** | **Scopus** |
| (microsimulation OR "synthetic population" OR "individual-based") AND (health OR disease OR wellbeing OR burden) AND ("climate change" OR environment OR weather OR "global warming" OR "climate crises") | 95 | 126 | 11 | 568 | 330 |

Table 1A. Search string and database search results (updated on 15 January 2025)

Table 2A. Methods summary (example)

| Climate Exposure | Health Outcome | Synthetic Population | Model methods | Exposure-response function and source | Projection and Time Steps | Citation |
|---|---|---|---|---|---|---|
| PM2.5 | Ischemic Heart Disease (IHD) incidence and mortality | 8.5M people in Greater London, disaggregated by age and gender. Age and gender structured directly from census data at the smallest geographical area. | **Dynamic spatial MSM**. The health state of each member of the population is determined annually through sequential Bernoulli trials, e.g. from non-IHD to IHD or death. GFR are applied to the female population age 15 - 44 to simulate newborns in the population. Social deprivation effects on mortality were simulated through a linear relationship between decile of a deprivation index and mortality risk. A cessation lag was implemented as health impacts are not assumed to occur immediately after PM2.5 reduction would occur. | Annual average air pollution exposures (from 2014) were mapped per LSOA area unit. RR of all-cause mortality and RR of IHD were obtained from literature and applied to the model to calculate the RR of PM2.5 exposure IHD incidence | Annually to 2050 | Symonds P, Hutchinson E, Ibbetson A, Taylor J, Milner J, Chalabi Z, Davies M, Wilkinson P. Sci Total Environ. 2019 Dec 20; 697:134105. https://doi.org/10.1016/j.scitotenv.2019.134105. |
